# Maternal and child outcomes reported by breastfeeding women following mRNA COVID-19 vaccination

**DOI:** 10.1101/2021.04.21.21255841

**Authors:** Kerri Bertrand, Gordon Honerkamp-Smith, Christina Chambers

## Abstract

**Table of Contents Summary:** *What’s known on this subject:* One published U.S. study with 31 breastfeeding women who were vaccinated with either of the two available mRNA COVID-19 vaccines found that 67% experienced any side effects after dose one and 61% after dose two of the vaccine.

*What this study adds:* More than 85% of 180 breastfeeding women who received an mRNA COVID-19 vaccine reported local or systemic symptoms, with higher frequency following the second dose. Few symptoms were reported in their breastfed children. No serious adverse events were noted.

*Contributors’ Statement Page:* Dr. Chambers and Ms. Bertrand designed the study and supervised the collection of data used in the study. Mr. Honerkamp-Smith performed the statistical analysis. All authors were involved in preparing the manuscript. All authors approved the final manuscript as submitted and agree to be accountable for all aspects of the work.

## INTRODUCTION

Clinical trials for the both the Pfizer-BioNTech BNT162b2 and Moderna mRNA-1273 COVID-19 vaccines demonstrated ability to prevent infection and severe disease, leading to emergency use authorization by the U.S. Food and Drug Administration.^1, 2^

The American College of Obstetrics and Gynecology and The Society for Maternal Fetal Medicine have recommended that these mRNA vaccines be made available for lactating women. However, initial trials excluded breastfeeding women, leading to questions about their safety.^3^ One study of 31 breastfeeding women who received an mRNA vaccine found that >60% reported side effects.^4^ We sought to evaluate a larger sample of vaccinated breastfeeding women and their breastfed children.

## MATERIALS AND METHODS

Breastfeeding women in the U.S. who received an mRNA vaccine were enrolled into the Mommy’s Milk Human Milk Research Biorepository at the University of California, San Diego. Demographics, health history, vaccine brand, and maternal symptoms and child events were collected by maternal interview and questionnaire for seven days following each dose. The study was approved by the UC San Diego IRB; written consent was obtained from participants.

Maternal and child characteristics and outcomes were compared by brand for each dose using Student’s t-test for continuous and Fisher’s exact test for categorical variables. Missing values were excluded. R version 25 4.0.3 was used for analyses.

## Results

Between December 14, 2020 and February 1, 2021, 180 women who received either mRNA vaccine were enrolled. As shown in Table 1, 128 women (71.1%) received the Pfizer vaccine and 52 (28.9%) received the Moderna brand. Child age at enrollment averaged 7.47 months (SD 5.44, Range 0.09 to 27.45 months). The majority (86.0%) were exclusively breastfed.

**Table 1:** Characteristics of breastfeeding women who received either mRNA COVID-19 vaccine and their breastfed children, N=180.

|  | **Pfizer/BioNTech**<br>**n=128** | **Moderna**<br>**n= 52** | **All**<br>**N= 180** |
| --- | --- | --- | --- |
| **Maternal Age (years)** | 34.92 (3.67) | 34.50 (3.24) | 34.80 (3.54) |
| **Race** |  |  |  |
| White | 119 (93.7) | 39 (75.0) | 159 (88.3)** |
| Black | 0 (0.0) | 1 (1.9) | 1 (0.6) |
| Asian | 7 (5.5) | 12 (23.1) | 19 (10.6) |
| Native American/ Alaska Native | 1 (0.8) | 0 (0.0) | 1 (0.6) |
| **Ethnicity** |  |  |  |
| Non-Hispanic | 123 (96.9) | 51 (98.1) | 174 (96.7) |
| Hispanic | 4 (3.1) | 1 (1.9) | 6 (3.3) |
| **Education** |  |  |  |
| Some College | 1 (0.8) | 0 (0.0) | 1 (0.6) |
| College Graduate | 33 (26.2) | 12 (23.5) | 45 (25.3) |
| Post-Graduate | 92 (73.0) | 39 (76.5) | 132 (74.2) |
| **Income per year** |  |  |  |
| \$10,001-\$49,999 | 0 (0.0) | 1 (2.0) | 1 (0.6) |
| >\$60,000 | 127 (100.0) | 50 (98.0) | 178 (99.4) |
| **Full Time Employment** | 89 (71.8) | 34 (66.7) | 124 (70.5) |
| **Maternal Co-Morbidities** |  |  |  |
| BMI >30 | 16 (12.6) | 4 (7.8) | 20 (11.2) |
| Hypertension | 2 (1.6) | 0 (0.0) | 2 (1.1) |
| Diabetes | 0 (0.0) | 0 (0.0) | 0 (0.0) |
| Asthma | 20 (15.7) | 6 (11.5) | 26 (14.4) |
| **Prior COVID-19 infection** | 4 (3.1) | 4 (7.7) | 8 (4.4) |
| **Child Age in months (SD, Range)** | 7.60 (5.54, 0.39-27.45) | 7.16 (5.26, 0.10-23.45) | 7.47 (5.44, 0.09 - 27.45) |
| **Child Sex** |  |  |  |
| Female | 75 (59.5) | 25 (49.0) | 100 (56.7) |
| Male | 51 (40.5) | 26 (51.0) | 77 (43.3) |
| **Pre-term Delivery** | 5 (3.9) | 4 (7.7) | 9 (5.0) |
| **Exclusive Breastfeeding** | 107 (84.3) | 45 (90.0) | 153 (86.0) |

\*p<0.05 \*\*p<0.005. Continuous variables were summarized by mean (SD) and group comparisons were made using a t-test. Categorical variables were summarized by count (%) and group comparisons were made using Fisher's Exact test.

Following dose one, similar proportions of women reported any vaccine symptom by brand (89.4% Pfizer; 98.1% Moderna); frequency by specific symptom did not differ by brand. However, following dose two, women who received the Moderna brand were significantly more likely to report systemic side effects including chills, muscle/body aches, fever, vomiting. They were also more likely to report localized symptoms including pain, redness, swelling or itching at the injection site than women following dose two of the Pfizer brand (all p’s <0.05). A small proportion of women following dose one of either vaccine reported a reduction in milk supply. There was a significant difference in reduction of milk supply following dose two by brand (8.0% vs. 23.4% for Pfizer and Moderna, respectively, p<0.05). However, in all cases milk production returned to normal within 72 hours. Three women reported a change in color of milk (blue-green) following dose one; two reported change in color of milk following dose two (Table 2).

**Table 2.**
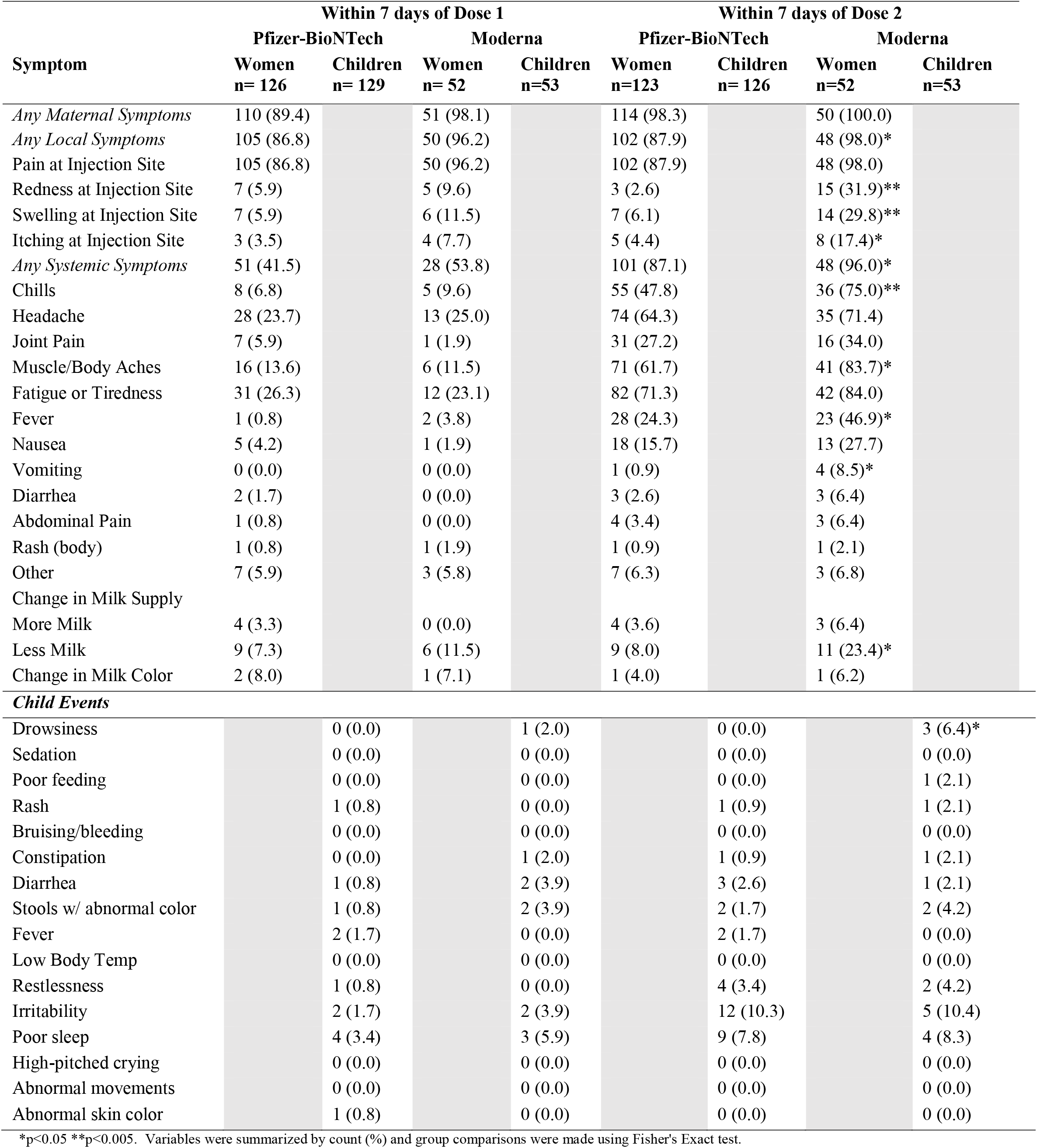
Symptoms reported by breastfeeding women who received either mRNA COVID-19 vaccine and events in their breastfed children.

Few events were reported for children following maternal vaccination with either brand or either dose. The most common child events following dose two were irritability (10.3% and 10.4% for Pfizer and Moderna, respectively), poor sleep (7.8% and 8.3% for Pfizer and Moderna respectively), with significantly more drowsiness reported for children whose mothers received Moderna vs. Pfizer (0% vs. 6.4%, p=0.02) (Table 2).

## Discussion

We found >85% of participants reported any symptoms for both the Pfizer-BioNTech and Moderna vaccines following either dose. This is substantially higher than the 61-67% of women who reported any symptom in the Gray et al. paper.^4^ This could be due to differences in methods of assessment, timing, and number of symptoms specifically queried. However, consistent with adult participants in clinical trials for each vaccine, we noted increased frequencies of most symptoms following the second dose compared to the first.^5, 6^ We also found significantly greater frequencies of localized pain, redness, swelling and itching at the injection site as well as systemic symptoms including chills, muscle/body aches, fever and vomiting following dose two of the Moderna brand vs. Pfizer. Some participants reported a reduction in milk supply, which in all cases returned to normal. We found low frequencies of any events reported in children, all of which were non-serious. To our knowledge there have been no previous studies published on outcomes in breastfed children.

These data are reassuring regarding the safety of vaccination in breastfeeding women and their breastfed children with either of the mRNA COVID-19 vaccines. Additional studies are underway to evaluate milk composition and antibody status in samples obtained from women participating in the current study.

## Data Availability

Data access may be provided, with appropriate ethics approval, by contacting the authors

## Acknowledgements

The authors wish to thank student researchers Angelique Ghadishah, Bianca Kermani, Samantha Powell, and Zoe Sidiropoulos for helping with data collection. We also wish to express our appreciation to the women who participated in this study for giving so generously of their time and effort to provide clinical information.

